# Predicting Musculoskeletal Adverse Events During Moderate- to High-Intensity Walking Training in Chronic Stroke

**DOI:** 10.64898/2026.04.16.26351040

**Authors:** Daria Pressler, Sarah M. Schwab-Farrell, Oluwole O. Awosika, Darcy S. Reisman, Sandra A. Billinger, Michael A. Riley, Pierce Boyne, the HIT-Stroke Trial investigators

## Abstract

**Background:** Moderate- to high-intensity walking training (M-HIT) is an established intervention for improving walking capacity in chronic stroke. Musculoskeletal (MSK) adverse events commonly occur during M-HIT, yet tools to identify individuals at higher risk are limited. Baseline clinical characteristics may provide insight into susceptibility to training-related MSK adverse events during M-HIT. Thus, this study aimed to develop and internally validate a model for predicting MSK adverse events during a 12-week M-HIT program in chronic stroke using baseline clinical characteristics.

**Methods:** Participants (n=100) from HIT-Stroke Trials 1 and 2 were included. Baseline clinical characteristics included measures of orthopedic history, pre-existing pain, motor function, recent exercise history, demographics and health characteristics, stroke chronicity, and psychological health. Logistic regression models evaluated all possible combinations of baseline characteristics with up to three predictors. Leave-one-out cross-validation was used for internal validation to mitigate overfitting. Predictive performance was quantified using the C-statistic, and the candidate model with the highest cross-validated C-statistic was selected as the final model.

**Results:** MSK adverse events occurred in 32.0% of participants. The optimal three-variable model included prior orthopedic condition (Odds ratio [OR] 3.02 [95% CI 1.14-8.64]), Fugl-Meyer lower extremity motor score (OR 1.14 [95% CI 1.02-1.28]), and self-reported participation in regular walking exercise (OR 0.17 [95% CI 0.05-0.49]) at baseline. This model demonstrated moderate discrimination (cross-validated C-statistic = 0.74; apparent C-statistic = 0.78).

**Conclusions:** Participants reporting at least one pre-existing lower extremity or lumbar spine orthopedic condition and those with better lower-extremity motor function exhibited greater odds of experiencing MSK adverse events during M-HIT, while participants reporting participation in regular walking exercise had lower odds. These findings suggest that baseline clinical characteristics may help identify individuals at elevated risk for MSK adverse events during M-HIT who may warrant closer monitoring or risk-reduction strategies. Future studies are needed for external validation.

**Clinical Trial Registration:** https://ClinicalTrials.gov; Unique identifiers: NCT03760016, NCT06268041

## INTRODUCTION

Moderate- to high-intensity walking training (M-HIT) is strongly recommended for individuals with chronic stroke^1,2^ and is increasingly emphasized in clinical rehabilitation programs targeting improvements in post-stroke walking function.^3–6^ M-HIT involves walking training performed at speeds exceeding an individual’s typical self-selected gait speed or at cardiovascular intensities exceeding 40% heart rate reserve.^2^ This approach has been shown to significantly improve walking speed and endurance (i.e., walking capacity) without appearing to increase the risk of *serious* adverse events (e.g., heart attack, recurrent stroke).^1,2,7–10^ Despite this favorable safety profile, treatment-related musculoskeletal (MSK) pain has been reported in approximately 30% of participants with stroke undergoing M-HIT.^2^

MSK adverse events, including treatment-related pain or injury involving the lower extremity or lumbar spine, are common across exercise- and therapy-based rehabilitation programs.^11–15^ Reported frequencies among individuals with stroke participating in M-HIT do not appear markedly higher than those observed in other stroke rehabilitation^2^ and exercise populations.^11–15^ However, MSK adverse events can still disrupt rehabilitation by necessitating reductions in training intensity or dosage and by diminishing treatment adherence, potentially limiting the effectiveness of M-HIT.^15–19^

Despite the clinical relevance of MSK adverse events, there is limited guidance to help clinicians anticipate which individuals are more likely to experience them during M-HIT. Improved ability to identify individuals at greater risk for MSK adverse events could support targeted risk-reduction strategies and individualized training progression during M-HIT. Prior research suggests that MSK pain and injury in stroke and other adult populations have been associated with baseline clinical characteristics across multiple clinical domains, including orthopedic history, pre-existing pain, motor function and mobility, recent exercise history, demographics and health characteristics, and psychological health.^20–34^ However, no studies have evaluated whether baseline clinical characteristics can differentiate individuals with chronic stroke who do versus do not experience MSK adverse events during M-HIT. Therefore, the purpose of this study was to develop and internally validate a clinical prediction model to identify individuals with chronic stroke who are more likely to experience MSK adverse events during M-HIT using baseline clinical characteristics.

## METHODS

### Data Availability

Data from HIT-Stroke Trial 1 are available in the NICHD Data and Specimen Hub (DASH): https://dash.nichd.nih.gov/study/424597 DOI: 10.57982/06fb-wh62. Data from HIT-Stroke Trial 2 will be available in DASH at the time of trial completion.

### Study Population

This retrospective observational analysis used data from the first 100 consecutively enrolled participants in HIT-Stroke Trials (HST) 1 and 2 between 2019 and 2025. HST 1 and 2 are randomized clinical trials evaluating the effectiveness of moderate-intensity continuous versus high-intensity interval walking training for improving walking capacity in individuals with chronic stroke.^10,35^ All participants were randomized to a standardized M-HIT protocol, receiving either moderate-intensity continuous training or high-intensity interval training, delivered as 45-minute sessions of combined overground and treadmill walking, 3 times per week for 12 weeks.

General eligibility criteria for HST, and therefore this study, included age 30-85 years, most recent stroke at least 6 months prior, walking speed <1.0 m/s, ability to walk 10 m overground without continuous assistance, ability to walk at least 3 minutes on the treadmill at ≥0.13 m/s (0.3 mph), stable cardiovascular condition, and ability to follow instructions and communicate with investigators. Participants were recruited from the community and provided written informed consent prior to participation. The study protocol was approved by the University of Cincinnati Institutional Review Board. This study followed the Transparent Reporting of a multivariable prediction model for Individual Prognosis or Diagnosis (TRIPOD) reporting guidelines.^36^ A completed TRIPOD checklist is provided in the Supplemental Materials.

### Baseline Data Collection

Clinical characteristics were collected prior to initiation of M-HIT. Participants completed standardized questionnaires that included assessments of medical history, demographic and stroke characteristics, pain history, aerobic exercise participation during the month prior to baseline assessment, and psychological health. Medical records were reviewed to supplement patient-reported medical history. Trained clinicians (e.g., physical and occupational therapists) performed standardized assessments of motor function and mobility, and height and weight were measured to calculate body mass index. From the available baseline data, 37 variables were selected *a priori* as candidate predictors based on prior evidence linking these characteristics to MSK pain and injury^20–34^ and their clinical relevance to walking training after stroke. For more detailed information on all candidate predictors, see Supplementary Table 1.

### Assessment of MSK Adverse Event Occurrence

The primary outcome of interest was the occurrence of at least one clinically relevant MSK adverse event involving the lower extremity and/or lumbar spine that was plausibly related to participation in M-HIT. More specifically, an MSK adverse event was defined as: (1) a musculoskeletal condition involving the lower extremity and/or lumbar spine, including overuse or acute injuries (e.g., strains, sprains) and exacerbations of pre-existing conditions (e.g., arthritis flare-ups); (2) occurrence during or within 24 hours of a training session; (3) absence of a more plausible alternative cause than M-HIT; and (4) clinical relevance, indicated by sustained increases in pain intensity (e.g., increase >2/10 persisting >48 hrs or increase >4/10 persisting >1 hr), interference with activities of daily living, modification or limitation of training dosage, observable increases in swelling or movement limitation, or the need for additional pain management or referral. Events attributable to non-MSK neurological symptoms (e.g., neuropathic pain, spasticity, fatigue, or tone changes), device or footwear irritation, falls as the primary mechanism, or upper-extremity conditions were excluded from this definition. Lower extremity and lumbar spine events were selected because they were deemed most likely to reflect MSK stress related to repetitive walking practice.

MSK adverse events were prospectively and systematically monitored during participation in the M-HIT protocol through continuous observations during sessions and structured participant interviews before and after each training session (Supplementary Table 2). All reported events were followed by clinical evaluation, including a physical assessment or referral when indicated. Adverse events were systematically documented and categorized by type, severity (grade 1-5), and causal relationship to the intervention. To ensure consistent, objective evaluation, all adverse events were initially adjudicated by a centralized blinded physician. For the purposes of this study, all adverse events were subsequently assessed by two additional blinded reviewers to independently determine whether each event met the predefined criteria for classification as an MSK adverse event as previously described. Agreement was high, with disagreements occurring in 6 of 151 adjudicated events (3.97%; 96% agreement). Disagreements in classification between the two independent reviewers were resolved through consensus discussion.

### Statistical Analysis

Logistic regression was used to develop a prediction model for MSK adverse event occurrence using baseline clinical characteristics. The ‘all possible regressions’ procedure^37^ was used to systematically evaluate all combinations of candidate baseline clinical predictors. When practical, this approach is preferred over any other variable selection strategy because it guarantees identification of the best-fitting model among the candidate predictor set.^37^ Model complexity was conservatively restricted to a maximum of 3 predictors to reduce overfitting and enhance model stability given the available sample size and anticipated event frequency (∼30%).^38^ Internal validation was performed using leave-one-out cross-validation (LOOCV) during model development to reduce optimism in performance estimates while maximizing use of the available data. Models were fit using complete-case data for the predictors included in each candidate model (i.e., participants with non-missing data for all predictors in that model). Missingness in baseline predictors was minimal (≤ 2% of participants). Given the limited extent of missing data and the primary focus on predictive model development and internal validation, this modified intent-to-treat approach was selected over multiple imputation.^38^

Model discrimination was assessed using the cross-validated C-statistic (area under the receiver operating characteristic [ROC] curve) calculated from the model predicted probability of MSK adverse events versus the observed event occurrence in the holdout data. The candidate model with the highest cross-validated C-statistic was selected as the final model. Bootstrap resampling (5,000 replicates) was used to estimate 95% confidence intervals (CIs) for apparent and cross-validated C-statistics. Model calibration reflects agreement between predicted and observed event probabilities. Calibration was assessed using the calibration intercept and slope calculated from cross-validated predicted probabilities,^39^ with 95% confidence intervals estimated using model-based Wald standard errors. Regression coefficients from the final model were exponentiated to obtain odds ratios, with 95% CIs derived using profile likelihood methods. Analyses were conducted in R (version 4.5.1) using the ‘pROC’ (version 1.19.0.1)^40^ and ‘CalibrationCurves’ (version 3.0.0)^41^ packages.

## RESULTS

### Participant Characteristics

The analytic sample included 100 participants, all of whom had complete MSK adverse event surveillance. Among these participants, 32 (32%) experienced at least one clinically relevant MSK adverse event plausibly related to M-HIT. Baseline clinical data were largely complete; however, two participants (one with and one without MSK adverse event occurrence) had partial missing data due to noncompletion of self-report questionnaire items. Consequently, sample sizes ranged from 98 to 100 across candidate models. Baseline participant characteristics stratified by MSK adverse event occurrence are presented in Table 1.

**Table 1.**
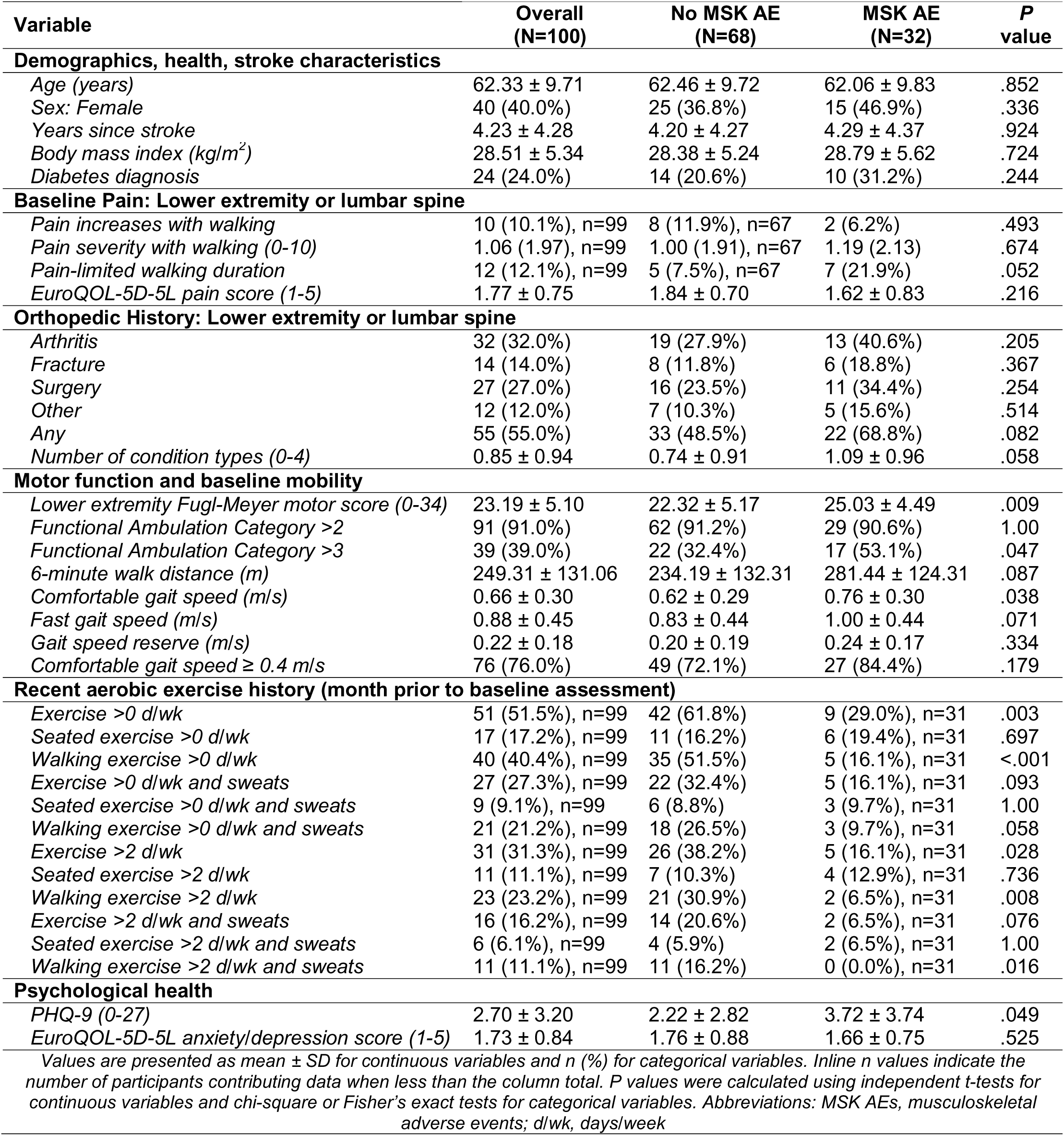
Baseline participant characteristics by MSK adverse event status.

|  |  |  |  |  |
| --- | --- | --- | --- | --- |
| Variable | Overall<br>(N=100) | No MSK AE<br>(N=68) | MSK AE<br>(N=32) | P<br>value |
| **Demographics, health, stroke characteristics** |  |  |  |  |
| Age (years) | 62.33 ± 9.71 | 62.46 ± 9.72 | 62.06 ± 9.83 | .852 |
| Sex: Female | 40 (40.0%) | 25 (36.8%) | 15 (46.9%) | .336 |
| Years since stroke | 4.23 ± 4.28 | 4.20 ± 4.27 | 4.29 ± 4.37 | .924 |
| Body mass index (kg/m<sup>2</sup>) | 28.51 ± 5.34 | 28.38 ± 5.24 | 28.79 ± 5.62 | .724 |
| Diabetes diagnosis | 24 (24.0%) | 14 (20.6%) | 10 (31.2%) | .244 |
| **Baseline Pain: Lower extremity or lumbar spine** |  |  |  |  |
| Pain increases with walking | 10 (10.1%), n=99 | 8 (11.9%), n=67 | 2 (6.2%) | .493 |
| Pain severity with walking (0-10) | 1.06 (1.97), n=99 | 1.00 (1.91), n=67 | 1.19 (2.13) | .674 |
| Pain-limited walking duration | 12 (12.1%), n=99 | 5 (7.5%), n=67 | 7 (21.9%) | .052 |
| EuroQOL-5D-5L pain score (1-5) | 1.77 ± 0.75 | 1.84 ± 0.70 | 1.62 ± 0.83 | .216 |
| **Orthopedic History: Lower extremity or lumbar spine** |  |  |  |  |
| Arthritis | 32 (32.0%) | 19 (27.9%) | 13 (40.6%) | .205 |
| Fracture | 14 (14.0%) | 8 (11.8%) | 6 (18.8%) | .367 |
| Surgery | 27 (27.0%) | 16 (23.5%) | 11 (34.4%) | .254 |
| Other | 12 (12.0%) | 7 (10.3%) | 5 (15.6%) | .514 |
| Any | 55 (55.0%) | 33 (48.5%) | 22 (68.8%) | .082 |
| Number of condition types (0-4) | 0.85 ± 0.94 | 0.74 ± 0.91 | 1.09 ± 0.96 | .058 |
| **Motor function and baseline mobility** |  |  |  |  |
| Lower extremity Fugl-Meyer motor score (0-34) | 23.19 ± 5.10 | 22.32 ± 5.17 | 25.03 ± 4.49 | .009 |
| Functional Ambulation Category >2 | 91 (91.0%) | 62 (91.2%) | 29 (90.6%) | 1.00 |
| Functional Ambulation Category >3 | 39 (39.0%) | 22 (32.4%) | 17 (53.1%) | .047 |
| 6-minute walk distance (m) | 249.31 ± 131.06 | 234.19 ± 132.31 | 281.44 ± 124.31 | .087 |
| Comfortable gait speed (m/s) | 0.66 ± 0.30 | 0.62 ± 0.29 | 0.76 ± 0.30 | .038 |
| Fast gait speed (m/s) | 0.88 ± 0.45 | 0.83 ± 0.44 | 1.00 ± 0.44 | .071 |
| Gait speed reserve (m/s) | 0.22 ± 0.18 | 0.20 ± 0.19 | 0.24 ± 0.17 | .334 |
| Comfortable gait speed ≥ 0.4 m/s | 76 (76.0%) | 49 (72.1%) | 27 (84.4%) | .179 |
| **Recent aerobic exercise history (month prior to baseline assessment)** |  |  |  |  |
| Exercise >0 d/wk | 51 (51.5%), n=99 | 42 (61.8%) | 9 (29.0%), n=31 | .003 |
| Seated exercise >0 d/wk | 17 (17.2%), n=99 | 11 (16.2%) | 6 (19.4%), n=31 | .697 |
| Walking exercise >0 d/wk | 40 (40.4%), n=99 | 35 (51.5%) | 5 (16.1%), n=31 | <.001 |
| Exercise >0 d/wk and sweats | 27 (27.3%), n=99 | 22 (32.4%) | 5 (16.1%), n=31 | .093 |
| Seated exercise >0 d/wk and sweats | 9 (9.1%), n=99 | 6 (8.8%) | 3 (9.7%), n=31 | 1.00 |
| Walking exercise >0 d/wk and sweats | 21 (21.2%), n=99 | 18 (26.5%) | 3 (9.7%), n=31 | .058 |
| Exercise >2 d/wk | 31 (31.3%), n=99 | 26 (38.2%) | 5 (16.1%), n=31 | .028 |
| Seated exercise >2 d/wk | 11 (11.1%), n=99 | 7 (10.3%) | 4 (12.9%), n=31 | .736 |
| Walking exercise >2 d/wk | 23 (23.2%), n=99 | 21 (30.9%) | 2 (6.5%), n=31 | .008 |
| Exercise >2 d/wk and sweats | 16 (16.2%), n=99 | 14 (20.6%) | 2 (6.5%), n=31 | .076 |
| Seated exercise >2 d/wk and sweats | 6 (6.1%), n=99 | 4 (5.9%) | 2 (6.5%), n=31 | 1.00 |
| Walking exercise >2 d/wk and sweats | 11 (11.1%), n=99 | 11 (16.2%) | 0 (0.0%), n=31 | .016 |
| **Psychological health** |  |  |  |  |
| PHQ-9 (0-27) | 2.70 ± 3.20 | 2.22 ± 2.82 | 3.72 ± 3.74 | .049 |
| EuroQOL-5D-5L anxiety/depression score (1-5) | 1.73 ± 0.84 | 1.76 ± 0.88 | 1.66 ± 0.75 | .525 |

Values are presented as mean ± SD for continuous variables and n (%) for categorical variables. Inline n values indicate the number of participants contributing data when less than the column total. P values were calculated using independent t-tests for continuous variables and chi-square or Fisher's exact tests for categorical variables. Abbreviations: MSK AEs, musculoskeletal adverse events; d/wk, days/week

### Final prediction model

The model with the highest cross-validated C-statistic included 3 baseline predictors and each one was statistically significant: history of prior lower extremity or lumbar spine orthopedic condition (*Any orthopedic condition*), lower extremity Fugl-Meyer motor score (LEFM), and self-reported participation in aerobic walking exercise >0 d/wk during the month prior to baseline assessment (Table 2). The top 10 models ranked by cross-validated C-statistic are summarized in Supplementary Table 3, with at least one predictor from the final model appearing in each of the top 10 models. Figure 1 and Table 3 show model-predicted probabilities of MSK adverse events across LEFM scores (0-34), stratified by orthopedic condition (yes/no) and regular walking exercise participation (yes/no).

**Figure 1.**
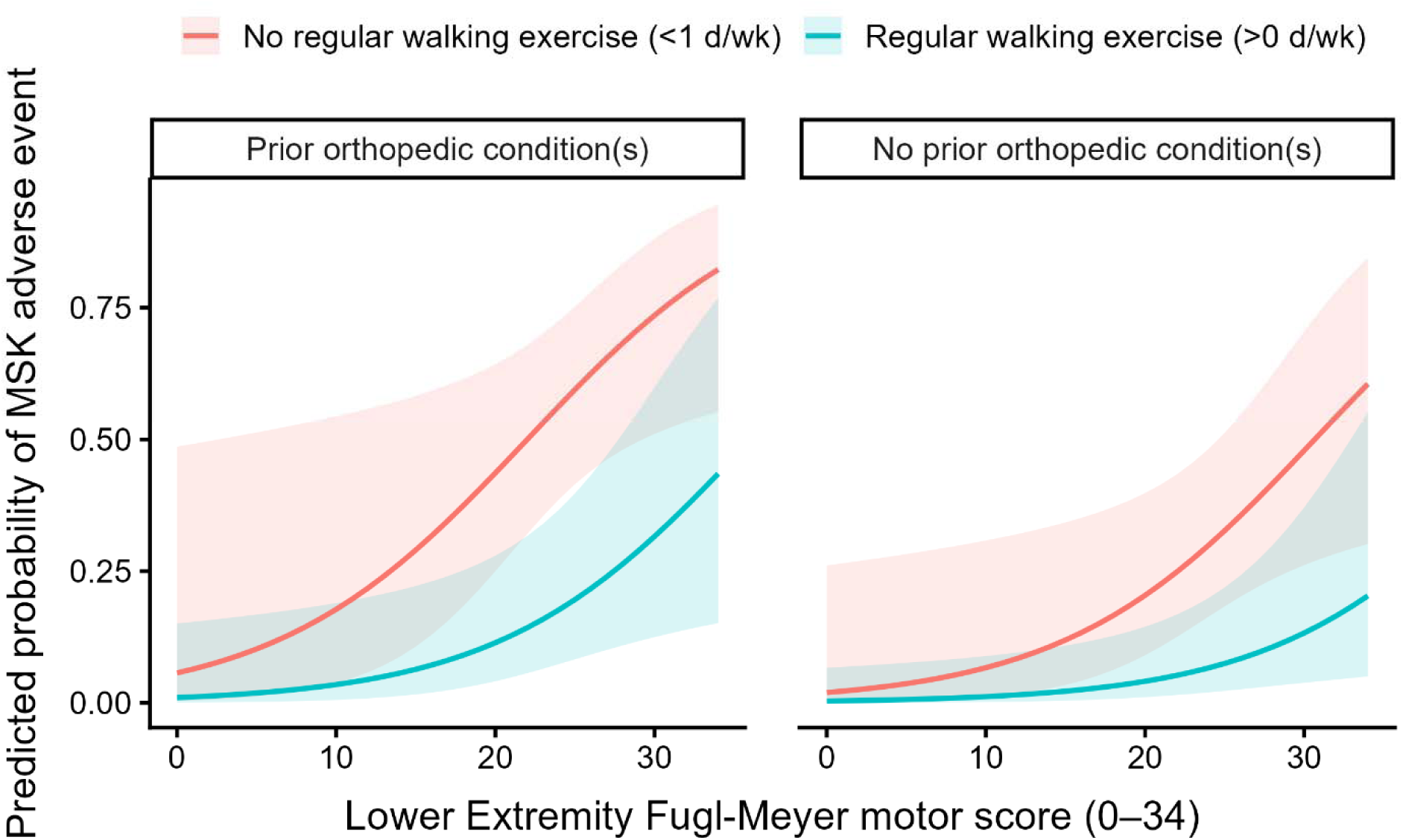
**Model-predicted probabilities of musculoskeletal (MSK) adverse events** across the lower extremity Fugl-Meyer motor score scale (0-34). Curves are shown separately by history of prior lower extremity and/or lumbar spine orthopedic condition (yes/no) and regular walking exercise (>0 d/wk) participation (yes/no). Shaded regions represent 95% confidence intervals.

**Table 2.**
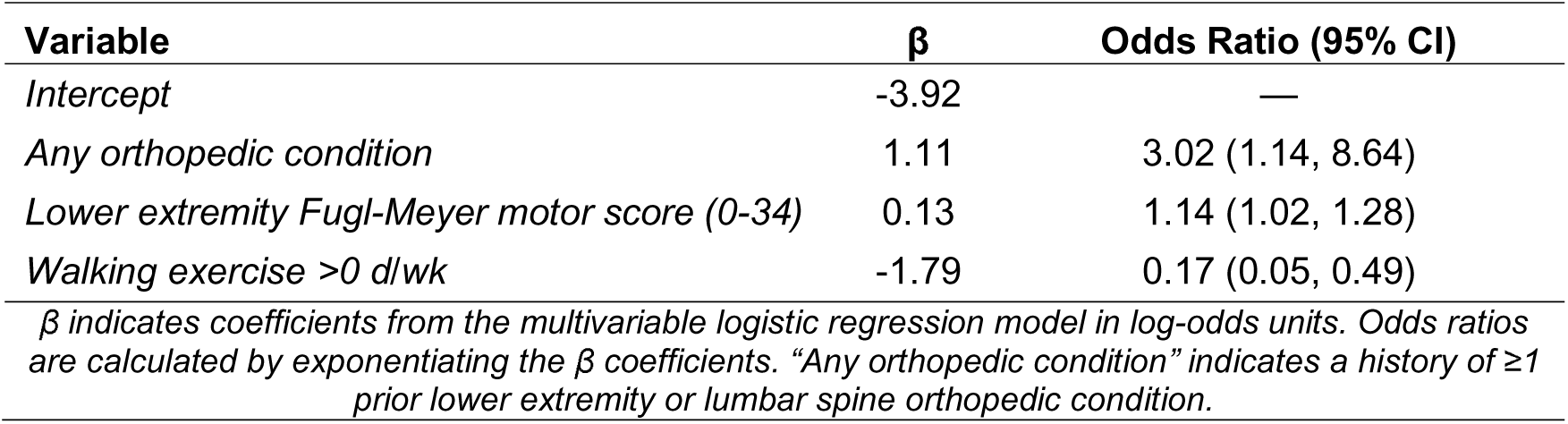
Regression coefficients and odds ratios for the final prediction model (n=99)

**Table 3.** Model-predicted probabilities of MSK adverse events.

| **Table 3. Model-predicted probabilities of MSK adverse events** |  |  |  |  |
| --- | --- | --- | --- | --- |
| **Orthopedic condition** | **Regular walking exercise >0 d/wk** | **LEFM = 9** | **LEFM = 24** | **LEFM = 34** |
| Yes | No | 0.16 (0.03, 0.54) | 0.56 (0.39, 0.73) | 0.82 (0.55, 0.95) |
| No | No | 0.06 (0.01, 0.30) | 0.30 (0.16, 0.48) | 0.61 (0.30, 0.84) |
| Yes | Yes | 0.03 (0.00, 0.18) | 0.18 (0.07, 0.37) | 0.43 (0.15, 0.77) |
| No | Yes | 0.01 (0.00, 0.09) | 0.07 (0.02, 0.20) | 0.20 (0.05, 0.55) |
| *Predicted probabilities (95% CI) derived from the final logistic regression model at LEFM scores of 9 (minimum observed), 24 (median), and 34 (maximum observed), stratified by orthopedic condition and regular walking exercise. Abbreviations: MSK, musculoskeletal; d/wk, days/week; LEFM, Lower Extremity Fugl-Meyer motor score* |  |  |  |  |

### Model Performance and Calibration

The final model demonstrated moderate discrimination, with a cross-validated C-statistic of 0.74 (95% CI, 0.63-0.83). Apparent model performance was higher (C-statistic = 0.78; 95% CI, 0.68-0.87), consistent with expected optimism. Cross-validated and apparent ROC curves are shown in Figure 2. Calibration was assessed using cross-validated predicted probabilities, yielding a calibration intercept near 0.00 (95% CI, -0.48-0.48) and a slope of 0.75 (95% CI, 0.62-0.83), suggesting minimal systematic prediction error with some evidence of overfitting. Visual inspection of the calibration plot demonstrated generally acceptable agreement between observed and cross-validated predicted event probabilities, with some overprediction at higher predicted probabilities (Figure 3).

**Figure 2.**
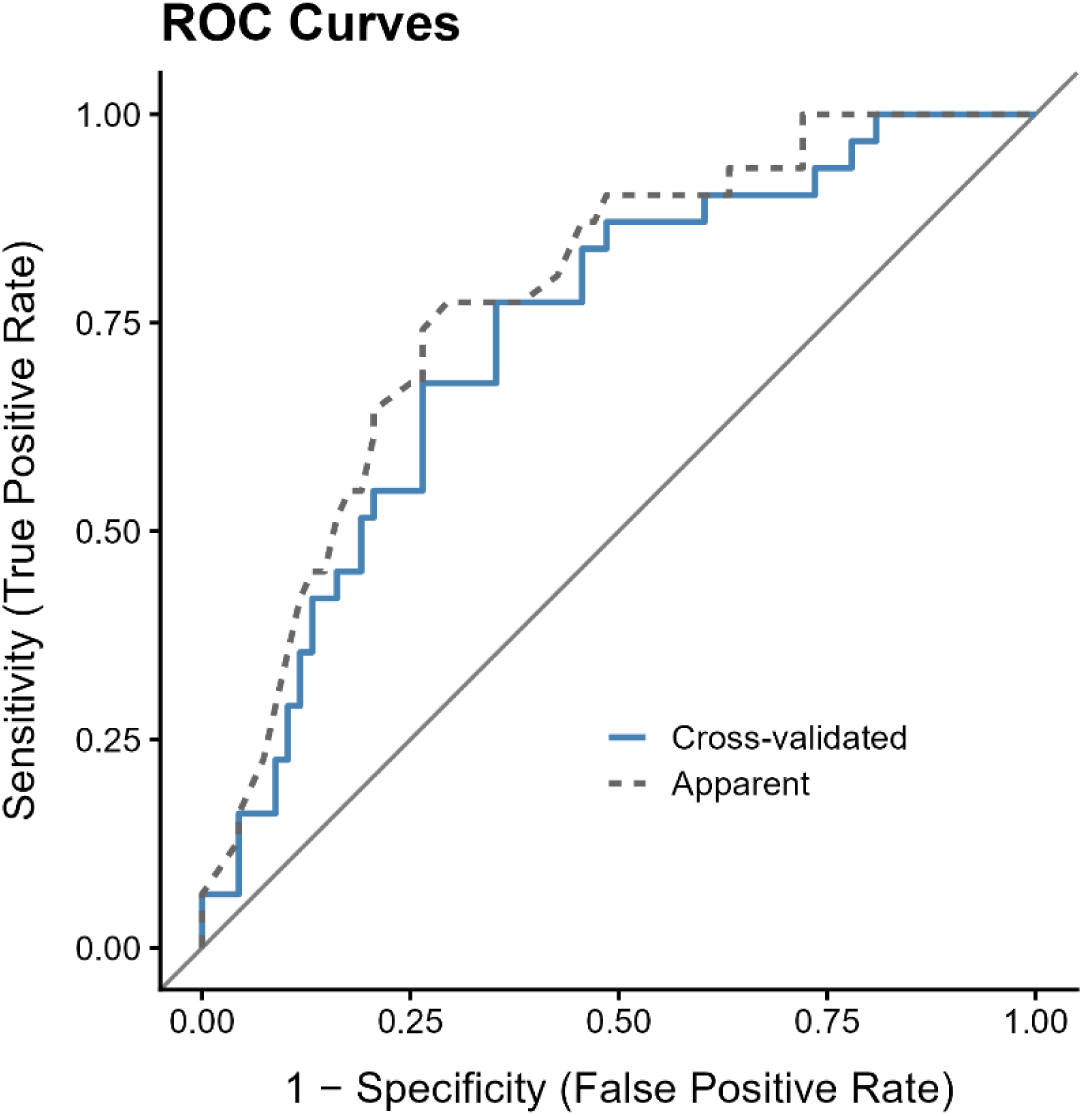
Receiver operating characteristic (ROC) curves for the final prediction model. Cross-validated and apparent ROC curves are shown.

**Figure 3.**
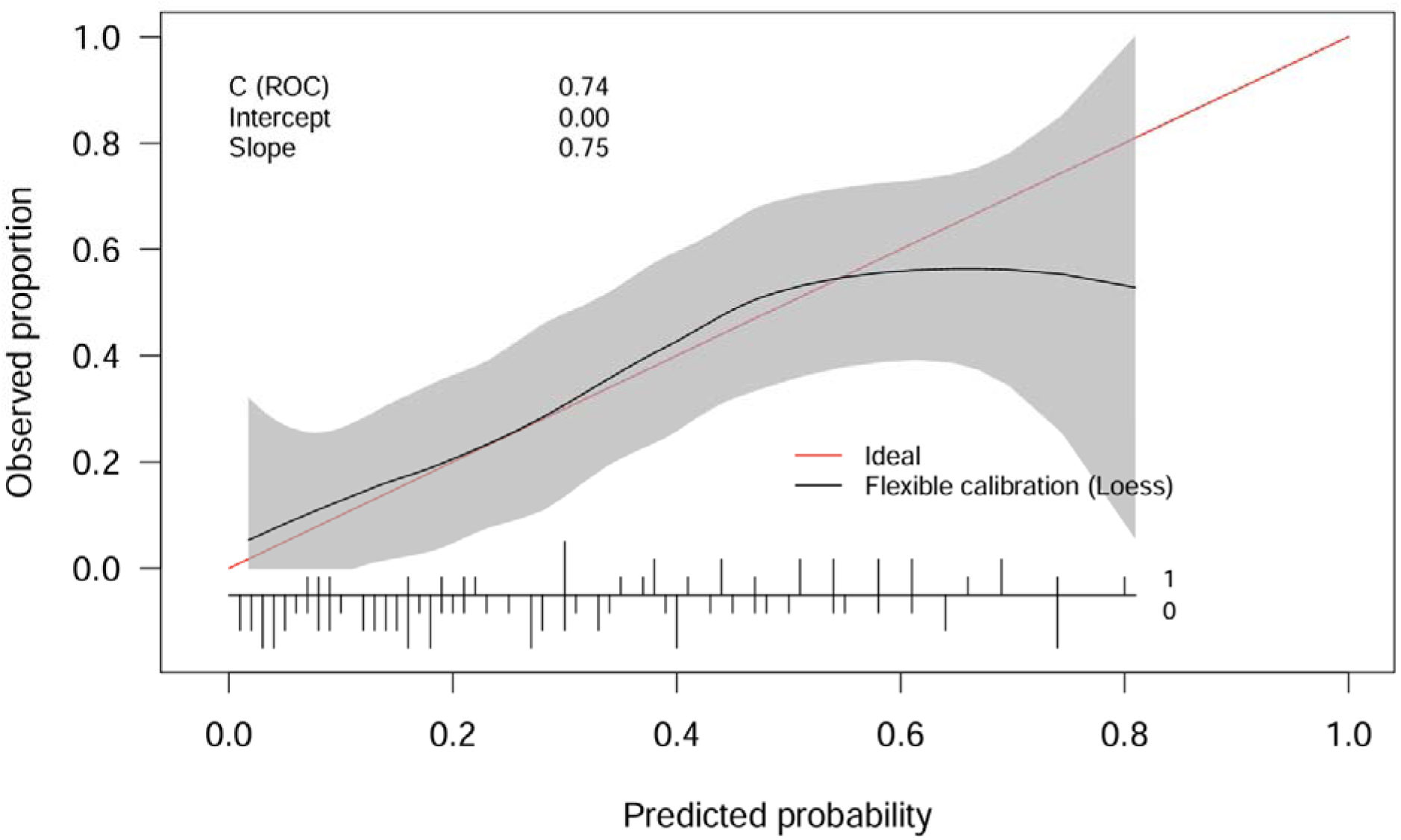
Cross-validated calibration. Calibration of the final prediction model, comparing observed and cross-validated predicted event probabilities.

## DISSCUSSION

### Final model predictors

In this study, we developed and internally validated a clinical prediction model to examine how commonly collected baseline clinical characteristics relate to the occurrence of MSK adverse events during M-HIT in individuals with chronic stroke. The final model included 3 statistically significant baseline clinical characteristics: history of at least one prior lower extremity and/or lumbar spine orthopedic condition, lower extremity motor impairment as measured by LEFM motor scores, and self-reported participation in aerobic walking exercise (>0 d/wk) during the month prior to baseline assessment.

Participants with a history of any lower extremity and/or lumbar spine orthopedic condition demonstrated significantly higher odds of experiencing a MSK adverse event during M-HIT (Table 2, Figure 1). This predictor encompassed a range of lumbar spine and lower extremity conditions, including degenerative disorders (e.g., osteoarthritis), prior fractures, prior surgeries, and other clinically relevant MSK conditions (e.g., osteoporosis). Pre-existing MSK pathology has similarly been identified as a risk marker for subsequent injury or symptom recurrence in physically active populations.^20–23^ Such pathology may also reflect persistent alterations in neuromuscular control, tissue capacity, or load distribution that could reduce tolerance to increased mechanical demand.^20,42^ In the context of M-HIT, prior orthopedic history may therefore represent a clinically accessible marker of increased susceptibility to MSK adverse events.

In contrast to the *positive* association between orthopedic conditions and MSK adverse events, participants who reported engaging in aerobic walking exercise (>0 days/week) within the month prior to baseline assessment demonstrated significantly lower odds of MSK adverse events (Table 2, Figure 1). This could be related to the load-management concept in athletic populations, which suggests that individuals with greater habitual exposure to mechanical loading may better tolerate subsequent increases in physical demand.^43,44^ In this context, even modest, self-selected walking performed prior to participation in M-HIT by individuals with chronic stroke may function protectively as a form of habitual loading that enhances tissue tolerance to repetitive mechanical walking stress. Alternatively, though perhaps not mutually exclusive, individuals who were more susceptible to MSK adverse events may have engaged in less walking-based exercise prior to participation. Thus, these potential patterns suggest that individuals with chronic stroke who previously engaged in walking exercise may be better positioned to tolerate the mechanical demands of M-HIT.

Interestingly, higher lower extremity motor function, as indicated by LEFM motor scores, was associated with significantly *increased* odds of MSK adverse events (Table 2). It seems unlikely that better motor function is an intrinsic risk factor for MSK adverse events. Rather, we suspect that individuals with greater baseline motor capacity may be able to achieve higher absolute walking speeds, greater step counts, or more rapid progression during M-HIT. Across repeated training sessions, this could translate to greater total mechanical loading exposure. Thus, we speculate that higher motor function may reflect increased opportunity for loading-related MSK adverse events rather than increased biological vulnerability.

### Model performance

The final model demonstrated moderate ability to distinguish between individuals who did and did not experience MSK adverse events during M-HIT. Model calibration was acceptable overall, although modest overprediction was observed at higher predicted risk levels. While this level of performance is unlikely to support definitive clinical decision-making at this stage, the model may provide a preliminary framework for identifying individuals who warrant closer monitoring of MSK symptoms during M-HIT. Collectively, these findings represent an initial step toward developing risk stratification tools for MSK adverse events during intensive locomotor rehabilitation.

### Limitations

This study represents an initial effort to develop a clinically interpretable prediction model for MSK adverse event occurrence during M-HIT in chronic stroke and should be interpreted within the appropriate context. The model was developed and internally validated within multi-center cohorts undergoing specific M-HIT protocols and has not yet undergone external validation. As such, findings may not generalize to individuals who do not meet the study eligibility criteria or to rehabilitation programs that differ in training intensity, frequency, or structure. Predictors were limited to baseline clinical characteristics collected in HST; incorporation of additional clinical, biomechanical, or session-level training variables may further refine risk estimation in future models. Moreover, MSK adverse events encompassed conditions affecting multiple anatomical regions that may arise through differing underlying mechanisms. Thus, future studies may improve prediction accuracy by modeling more specific MSK adverse event subtypes.

## Conclusions

Participants reporting at least one pre-existing lower extremity or lumbar spine orthopedic condition and those with better lower-extremity motor function demonstrated greater odds of MSK adverse events during M-HIT, whereas individuals reporting regular walking exercise (>0 days/week) at baseline demonstrated lower odds. Collectively, these predictors suggest that MSK adverse events during M-HIT in chronic stroke are multifactorial and may reflect the combined influence of baseline MSK status, habitual loading exposure, and neurological capacity rather than any single domain in isolation. Importantly, these baseline clinical characteristics are readily obtainable in clinical practice and may provide a practical means of identifying individuals who may warrant closer monitoring of MSK symptoms during M-HIT. Future studies are needed for external validation.

## Supporting information

TRIPOD Reporting Checklist

## ACKNOWLEDGEMENTS

The authors thank the participants and research teams of HIT-Stroke Trials 1 and 2 for their contributions to this research.

## SUPPLEMENTAL MATERIALS

Tables S1-S3

TRIPOD Reporting Checklist

## APPENDIX

### HIT-Stroke Trial Investigators

Bria L. Bartsch; Amanda Briton-Carpenter; Sofia Buckley; Jamiah Burson; Daniel Carl; Matthew DeLange; Sarah Doren; Kari Dunning; Melinda Earnest; Amanda Engler; Jolene Foster; Christina Garrity; Myron Gerson; Madison Henry; Alli Horning; Jemma Kim; Brett M. Kissela; Jane C. Khoury; Abigail Laughlin; Kiersten McCartney; Thomas McQuaid; Allison Miller; Alexandra Moores; Jacqueline A. Palmer; Heidi Sucharew; Elizabeth D. Thompson; Katherine Walters; Jaimie Ward; Emily Wasik; Alicen A. Whitaker; Henry Wright; Erin Wagner; Madison Yeazell

