## Supplemental Tables S1-S3 for "Predicting Musculoskeletal Adverse Events During Moderate- to High-Intensity Walking Training in Chronic Stroke"

| Table S1. Candidate clinical predictors considered for model development | |
| --- | --- |
| Construct | **Variables** |
| Baseline Pain | *Walking-related pain: *Participants reporting lower extremity or lumbar spine pain during walking were asked: “Does pain limit how far or how long you can walk?” (yes/no), “How severe is the pain typically while walking?” (0-10), and “Does the pain increase the longer you walk?” (yes/no).* |
|  | EuroQOL-5D-5L Pain item: *Scale ranges from 1 (no pain) to 5 (extreme pain)* |
| Orthopedic History | Presence of a chronic lower extremity or lumbar spine MSK condition (e.g., arthritis) |
|  | History of lower extremity or lumbar spine orthopedic surgery (e.g., joint arthroplasty) |
|  | History of lower extremity or lumbar spine fracture |
|  | Other lower extremity or lumbar spine orthopedic problems (e.g., known osteoporosis or osteopenia) |
|  | History of any of the above lower extremity or lumbar spine orthopedic conditions |
|  | Number of orthopedic conditions (0-4): *Tallied from the above 4 orthopedic conditions,* |
| Motor Function & Baseline Mobility | Lower Extremity Fugl-Meyer: *Lower score indicates more severe motor function impairment* |
|  | Functional Ambulation Category (>2 or >3): *Higher score indicates greater independence* |
|  | Six-minute walk test distance (m) |
|  | Comfortable gait speed (m/s) |
|  | Fast gait speed (m/s) |
|  | Gait speed reserve: Fast – comfortable gait speed (m/s) |
|  | Comfortable gait speed ≥ 0.4 m/s (yes/no) |
| Recent Aerobic Exercise History | *^†^*Recent aerobic exercise exposure: *Participants were asked how often they performed aerobic exercise in the past month (None, < 1 time per week, 1-2 times per week, 3-5 times per week, > 5 times per week), how the exercise was performed (seated and/or walking), and if the exercise was vigorous enough to make them sweat (yes/no).* |
| Demographics, Health, & Stroke Characteristics | Demographics: *Age, sex* |
|  | Health status: *Body Mass Index (BMI), Diabetes diagnosis* |
|  | Stroke Chronicity: *Time since stroke* |
| Psychological Health | Patient Health Questionnaire-9: *Measure of depression (higher score indicates greater severity)* |
|  | EuroQOL-5D-5L Depression/Anxiety item: *Scale ranges from 1 (not anxious or depressed) to 5 (extremely anxious or depressed)* |
| **Responses were used to derive pain-related candidates for model development.*  *^†^Responses were used to derive exercise-related candidates of exercise frequency, mode, and intensity for model development.* | |

| Table S2. MSK adverse event structured patient interview questions | | | | |
| --- | --- | --- | --- | --- |
|  | **Questions (every visit)** | **Response Options** | **Follow-up Trigger** | **Follow-up Question** |
| Pre-session | *Did you have soreness from your last study visit?* | Yes / No | If Yes | Did it last >48 hours or interfere with iADL? *“How long did it last? Did it stop you from doing your normal daily activities?”* (If yes, start new adverse event form) |
| Pre-session | *Did you have a significant increase in your overall fatigue level from the last study visit?* | Yes / No | If Yes | Did it last >48 hours or interfere with iADL? *“How long did it last? Did it stop you from doing your normal daily activities?”* (If yes, start new adverse event form) |
| Pre-session | *Since the last study visit, have you had any falls or injuries?* | Yes / No | If Yes | Start a new Adverse Event form. |
| Pre-session | *Any medication changes?* | Yes / No | If Yes | Specify Medication changes below and update medication form(s) from initial screening visit |
| Pre-session | *Are you having any pain right now?* | Yes / No | If Yes | *How severe is the pain, if 0 is no pain and 10 is the worst pain you can imagine?* |
| Pre-session | *Have you had any changes to your health since the last study visit?* | Yes / No | If Yes | May need to start a new adverse event form |
| Post-session | *Any falls or injuries during the visit? (not including falls caught by therapist or harness unless participant was injured)* | Yes / No | If Yes | Start new adverse event form |
| Post-session | *Any need for assistance from therapist or harness to prevent fall or injury during visit?* | Yes / No |  |  |
| Post-session | *“Are you having any pain right now?”* | Yes / No | If Yes | *“How severe is the pain, if 0 is no pain and 10 is the worst pain you can imagine?”* |
| Post-session | *Is this pain new or significantly worse since the beginning of the visit?* | Yes / No | If Yes | Start new adverse event form |
| Post-session | *Did you have any lightheadedness or nausea during the study visit?* | Yes / No | If Yes | Start new adverse event form |
| Post-session | *Any other adverse events during visit?* | Yes / No | If Yes | Start new adverse event form |
| *Pre/post-visit questions pertaining to active screening for all types of adverse events. MSK adverse events were determined specifically from adverse event reporting based on criteria stated in main text.* | | | | |

| Table S3. Top 10 candidate models ranked by cross-validated c-statistic | | | |
| --- | --- | --- | --- |
|  | **Model** | n | C-Statistic |
| 1 | ***Orthopedic condition; LEFM motor score (0-34)****;* ***Regular walking exercise (>0 d/wk)*** | 99 | 0.735 |
| 2 | *Pain-limited walking duration;* ***LEFM motor score (0-34)****;* ***Regular walking exercise (>0 d/wk)*** | 98 | 0.730 |
| 3 | ***Orthopedic condition****;* *6-minute walk distance (m);* ***Regular walking exercise (>0 d/wk)*** | 99 | 0.729 |
| 4 | ***Orthopedic condition****; Comfortable gait speed (m/s);* ***Regular walking exercise (>0 d/wk)*** | 99 | 0.726 |
| 5 | ***Orthopedic condition****;* ***Regular walking exercise (>0 d/wk)****; PHQ-9 (0-27)* | 99 | 0.725 |
| 6 | ***Orthopedic condition****; Fast gait speed (m/s);* ***Regular walking exercise (>0 d/wk)*** | 99 | 0.723 |
| 7 | *6-minute walk distance (m);* ***Regular walking exercise (>0 d/wk)****;* *PHQ-9 (0-27)* | 99 | 0.722 |
| 8 | ***LEFM motor score (0-34)****;* ***Regular walking exercise (>0 d/wk)****; Frequent walking exercise + Sweat (>2 d/wk + sweating)* | 99 | 0.720 |
| 9 | *Comfortable gait speed (m/s);* ***Regular walking exercise (>0 d/wk)****; PHQ-9 (0-27)* | 99 | 0.720 |
| 10 | ***Orthopedic condition****; Functional Ambulation Category >3;* ***Regular walking exercise (>0 d/wk)*** | 99 | 0.719 |
| *Models were ranked by cross-validated C-statistic. All models were limited to maximum of three predictors and evaluated using leave-one-out cross-validation. Bolded predictors indicate variables included in the final model. Direction and magnitude of associations for the final selected model are reported in Table 2*  *Abbreviations: LEFM, Lower extremity Fugl-Meyer* | | | |
